# How does the weekend catch-up sleep ratio affect the health and lifestyle of Korean adults? An age- and gender- matched study

**DOI:** 10.1101/2023.09.04.23295027

**Authors:** In-Whi Hwang, Soo-Ji Hwang, Jun-Hao Shen, Jisu Kim, Jung-Min Lee

## Abstract

The purpose of this study was to investigate the association between various levels of physical activity, self-perception, cardiometabolic risk factors, and weekend catch-up sleep ratio (CSR). Using raw data from the Korea National Health and Nutrition Examination Survey 2018 – 2021, all participants were divided into three groups (< 1.0, 1.0 ≤ CSR < 1.5, or ≥ 1.5) by CSR, which is the value calculated by dividing weekend sleep time by weekday sleep time. After matching age and gender, 2,484 Korean adults were selected as study participants. Descriptive statistics, chi-square test, ANOVA, and multinomial logistic regression analysis were performed to analyze the data. The results showed a significant association between weekend CSR and socioeconomic status, physical activity, self-perception levels, and cardiometabolic risk factors. Specifically, compared to the reference group (1 ≤ CSR < 1.5), those with a CSR < 1 were 2.42 times more likely to live in a single-family house (OR = 2.42, 95% CI = 1.77 – 3.32) and 1.71 times more likely to engage in vigorous physical activity meeting WHO guidelines (OR = 1.71, 95% CI = 1.20 – 2.46). They were also 1.33 times more likely to perceive themselves as ‘obese’ (OR = 1.33, 95% CI = 1.00 – 1.76). Conversely, those in the CSR ≥ 1.5 group were 3.93 times more likely to be ‘pink-collar’ workers (OR = 3.93, 95% CI = 2.70 – 5.71), 1.72 times more likely to perceive their stress levels as ‘quite’ (OR = 1.72, 95% CI = 1.20 – 2.47), and 1.87 times more likely to have diabetes (OR = 1.87, 95% CI = 0.97 – 3.60). Alterations in CSR could indicate changes in physical activity levels, sedentary behavior duration, and other health indicators, ultimately influencing overall well-being. Therefore, a comprehensive healthcare approach incorporating CSR considerations is increasingly essential.

## Introduction

The global prevalence of sleep disorders and sleep deprivation has been steadily increasing, with factors such as modern lifestyle, work culture, and increased screen time contributing to this concerning trend (1-3). South Korea is no exception, experiencing a growing number of individuals reporting sleep deprivation (4). The concept of catch-up sleep, which refers to the compensatory sleep obtained during weekends or days off, has emerged as a typical response to sleep debt accumulated during weekdays due to demanding work or social obligations (5). This phenomenon is prevalent among Koreans, as they grapple with long working hours, high-stress lifestyles, and societal expectations that often lead to sleep deprivation and irregular sleep patterns (6-8).

Previous studies revealed that physical activity (PA) is essential to maintaining overall health and well-being (9, 10). Regular engagement in PA can improve cardiovascular health, reduce the risk of developing chronic diseases, enhance mental health, and promote better sleep quality (11-14). As the general awareness of the importance of regular participation in PA, it is crucial to understand the factors that may influence an individual’s level of PA, such as sleep deprivation (15, 16). However, there is insufficient research to confirm how the weekend catch-up sleep ratio (CSR) impacts PA levels among Korean adults.

Sedentary behavior (SB), defined by extended periods of inactivity or sitting, has become increasingly prevalent in modern society, contributing to various health problems such as sleep deprivation, obesity, type 2 diabetes, and cardiometabolic diseases (17-20). SB is becoming a significant contributor to mortality, particularly with the rise of sedentary occupations and technology-driven lifestyles (21). Despite achieving sufficient levels of PA, the continuous link between SB and the onset and progression of chronic conditions has been confirmed by a study by Biswas and colleagues, who demonstrated that extended periods of inactivity are independently associated with negative health outcomes, irrespective of PA levels (22). This changing paradigm of modern lifestyles has increased in SB time, characterized by prolonged inactivity and irregular sleep patterns (23).

Our mental health, deeply influenced by stress and subjective perception, significantly impacts sleep quality (24, 25). The persistent demands of daily life often generate considerable stress, which can precipitate mental health disorders like anxiety and depression (26). Simultaneously, subjective perception of body shape influenced by societal standards can contribute to mental health problems, potentially leading to disorders such as body dysmorphia or eating disorders (27). Additionally, a negative self-evaluation of health, such as an exaggerated belief in personal physical decline or illness, can increase stress levels, induce anxiety, and disrupt normal sleep, contributing to an unstable CSR (28, 29). Therefore, effective personal management strategies are crucial to address the relationship between mental health and sleep patterns influenced by these factors.

Cardiometabolic risk factors (CRF), such as obesity, hypertension, dyslipidemia, and insulin resistance, have been associated with sleep deprivation in numerous studies (30-32). This connection involves hormonal imbalances, impaired glucose metabolism, and increased inflammation (33, 34). Sleep deprivation can exacerbate existing cardiometabolic issues and potentially lead to new health problems (35). Besides directly impacting physical health, poor sleep also profoundly affects mental health (36). It can lead to elevated stress and anxiety levels, further complicating the management of cardiometabolic health (37). Thus, understanding and addressing sleep deprivation is crucial for cardiometabolic health and overall well-being (38).

Previous research has primarily focused on the association between sleep deprivation and various health-related variables. However, these studies have not extensively explored the specific role of CSR in PA, SB, mental health, and CRF among Korean adults. The proposed research aims to address this gap by examining the impact of CSR on these health-related factors. This research will provide a more comprehensive understanding of the complex associations between CSR and health-related factors and identify potential confounding factors such as socioeconomic status and lifestyle habits. Ultimately, the findings from this study will contribute valuable insights that can inform the development of tailored interventions and public health strategies for the Korean adult population.

## Methods

### Study participants

This cross-sectional study used data from the Korea National Health and Nutrition Examination Survey (KNHANES) during 2018 – 2021. The KNHANES is a comprehensive national survey implemented across South Korea. Its primary objective is to consistently accumulate data annually encompassing various elements, encompassing sociodemographic indicators, economic status, and health-related characteristics and behaviors across all age demographics. Since 2007, this body of data has undergone review and received an annual endorsement by the Research Ethics Review Committee from the Korea Centers for Disease Control and Prevention Agency (KDCA). The KNHANES data is an open-access resource for scholars, enabling further exploratory analysis and research in numerous fields.

The KNHANES for 2018 – 2021 was conducted with 30,551 respondents. However, 5,616 individuals under 20 years were excluded from the data to align with the research parameters. In addition, participants who did not respond to the critical variables under examination were also eliminated from consideration. The remaining participants were classified into three distinct groups according to their CSR (CSR < 1, 1 ≤ CSR < 1.5, 1.5 ≤ CSR). These groups contained 1,112, 18,743, and 1,233 participants, respectively. Then, we matched participants based on age and gender, using the group with a CSR < 1 as the reference group. As a result of this stringent selection process, a balanced cohort of 828 participants from each group was extracted, resulting in a total study sample of 2,848 individuals.

### Group

The group was divided by considering the catch-up sleep ratio.

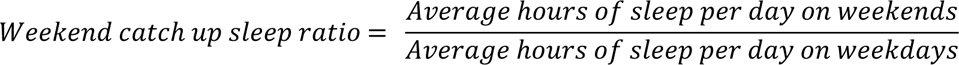

Using this formula, we classified the participants into three distinct groups according to their CSR (CSR < 1, 1 ≤ CSR < 1.5, 1.5 ≤ CSR). For instance, if participants sleep the same amount of time on weekdays and weekends, they are classified into a 1 ≤ CSR < 1.5 group. If a participant sleeps for 6 hours on weekdays and catches up by sleeping for 10 hours on weekends, their CSR is 1.5 or greater, placing them in the group with a 1.5 ≤ CSR group. Conversely, if participants sleep less on weekdays than on weekends, they belong to the CSR < 1 group.

According to prevailing health guidelines, the recommended sleep duration for adults typically falls within the range of 7 to 9 hours (39). However, it’s been observed that Korean adults tend to sleep less, with their average sleep duration falling below 6.5 hours (40). This discrepancy can be primarily attributed to socio-cultural norms and professional obligations within South Korea, which may significantly influence sleep behaviors, often encouraging a tendency to compensate by sleeping longer on weekends. Considering these factors, a normative threshold has been proposed in which weekend sleep duration should not surpass 1.50 times the corresponding weekday sleep duration to be deemed within acceptable bounds.

### Measurement

#### Anthropometric

KNHANES gathers data on gender and age via self-reported surveys. This study’s age variable was segmented into decade intervals ranging from the 20s to the 70s or above for further analysis. Anthropometric assessments were carried out by well-trained examiners adhering to standard protocols. The height (m2) measurement was performed using a stadiometer with participants standing erect, shoeless, and with heels, buttocks, and shoulders in contact with the apparatus. Body weight (kg) was measured using a digital scale, with participants dressed in light and barefooted attire. The waist circumference (WC, cm) was determined at the mid-point between the lower rib margin and the iliac crest using an inelastic measuring tape. Body Mass Index (BMI, kg/m2) was calculated as the weight divided by the square of the height, and participants were categorized as underweight, normal weight, overweight, or obese based on the World Health Organization (WHO) guidelines (World Health Organization).

### Socioeconomic status

The socioeconomic factors included type of residence: (1) single-family house, (2) apartment, (3) others, occupational status: (1) white-collar, (2) pink-collar, (3) blue-collar, (4) not classified, work schedule: (1) day, (2) night, (3) shift work (others), and household personal income quintile: (1) low, (2) lower middle, (3) middle, (4) upper middle, (5) high.

#### Lifestyle behaviors

The lifestyle factors included drinking status and smoking status. For drinking status in the KNHANES, participants were classified as: (1) abstained in the last year, (2) less than once a month, (3) about once a month, (4) 2 – 4 times a month, (5) 2 – 3 times a week, (6) 4 or more times a week, but considering the number of people belonging to each variable, we divided it into (1) abstained in the last year, (2) once a month, (3) 2 – 4 times a month, (4) at least twice a week. Smoking status was categorized as (1) current smoker, (2) ex-smoker, (3) never smoked.

#### Health-related factors

The health-related factors included in this study encompass moderate to vigorous physical activity (MVPA) participation time, the number of days walking per week, SB time, perceived self-body shape, perceived self-health status, perceived stress level, average daily sleep duration (weekday, weekend), and CRF. The time spent on MVPA was calculated using the following process, in line with the WHO’s recommendations for PA (World Health Organization, 2022). Participants were asked about time on vigorous PA during their work or leisure, resulting in shortness of breath or a speedy heart rate. The daily hours and minutes spent on these activities were added and multiplied by the number of days a week. This resulted in the total weekly hours of vigorous PA. A similar process was used to calculate the time spent on moderate PA, including work, leisure activities, or moving from place to place that causes slight shortness of breath or a slightly rapid heart rate. According to WHO’s recommendation, we separated the groups into (1) met and (2) not met for MVPA variables. Considering the distribution of participants, responses regarding the number of days walked per week have been categorized into (1) every day, (2) 4 – 6 days, (3) 1 – 3 days, and (4) not participating. For the analysis of SB time, we divided participants into groups based on the total daily duration of their SB time. We calculated the average daily sitting time in minutes for each participant and then categorized them into tertiles: (1) SB time < 420, (2) 420 ≤ SB time < 600, (3) 600 ≤ SB time. Sleep duration was assessed via a self-reported questionnaire, differentiating the average daily sleep duration (mins) into weekdays and weekends.

Cognition-related variables such as perceived self-body shape, perceived self-health status, and perceived stress level were classified through a self-report questionnaire given to the participants. The questions included ‘What do you think of your current body shape?’, ‘How do you feel about your health in general?’, and ‘How much stress do you feel daily?’. Responses regarding body shape were categorized as (1) thin, (2) average, and (3) fat. Health status responses were classified as (1) good, (2) average, and (3) poor. Lastly, perceived stress levels were separated into (1) not at all, (2) a little, and (3) quite.

The researchers of the KNHANES study procured explicit written authorization from the participants after delivering a comprehensive explanation of the objectives and methodology of CRF, encompassing measurements for obesity, hypertension, fasting glucose, total cholesterol, HDL cholesterol, triglycerides, and HbA1c. Participants were counseled to observe a minimum fasting period of 8 hours before the blood draw to ensure precise quantification of these health biomarkers. The resulting data were stratified into distinct categories according to predefined criteria established by the WHO. Obesity; (1) underweight (BMI < 18.5 kg/m2), (2) normal (18.5 kg/m2 ≤ BMI < 23 kg/m2), (3) overweight (23 kg/m2 ≤ BMI < 25 kg/m2), (4) obese (25 kg/m2 ≤ BMI). Hypertension; (1) normal (systolic < 120 mmHg and diastolic < 80 mmHg), (2) prehypertension (systolic 120 – 139 mmHg or diastolic 80 – 89 mmHg), and (3) hypertension (systolic ≥ 140 mmHg or diastolic ≥ 90 mmHg), Fasting glucose; (1) normal (70 – 99 mg/dL), (2) impaired fasting glucose (100 – 125 mg/dL), (3) diabetes (≥ 126 mg/dL), Total cholesterol; (1) normal (< 200 mg/dL), (2) borderline (200 – 239 mg/dL), and (3) hyperlipidemia (≥ 240 mg/dL), HDL cholesterol; (1) low (< 40 mg/dL), (2) normal (40 – 59 mg/dL), and (3) high (≥ 60 mg/dL), Triglycerides; (1) normal (< 150 mg/dL), (2) borderline (150 – 199 mg/dL), and (3) high (≥ 200 mg/dL), HbA1c; (1) normal (< 5.7%), (2) borderline (5.7 – 6.4%), and (3) diabetes (≥ 6.5%).

### Data Analysis

All data processing and statistical analysis used SPSS 28.0 version (SPSS Inc., Chicago, IL, USA). Descriptive statistics were conducted to summarize the participants’ characteristics for the anthropometric variables (gender, age, height, weight, WC, and BMI). The chi-square analysis (χ2 test) was used to compare the categorical data of general characteristics between the three groups. An analysis of variance (ANOVA) was conducted to analyze the mean difference of continuous variables (i.e., MVPA time, SB time, sleep duration, and work time) between the three groups. Following the ANOVA, a Bonferroni posthoc test was conducted to provide a more detailed analysis of the differences between the three groups. Moreover, multinomial logistic regression analysis was performed to find out the associations between the factors and CSR. Results are presented as odds ratios (OR) with 95% confidence intervals (95% CI), and the statistical significance of all analyzes was set by p < 0.05.

## Results

Table 1 presents the characteristics and anthropometric data of participants (n = 2,484) across three groups classified based on the CSR: CSR < 1 (n = 828), 1 ≤ CSR < 1.5 (n = 828), and 1.5 ≤ CSR (n = 828). The distribution of age and gender was identical across all groups due to the matching procedure conducted in the study. Regarding anthropometrics, male and female participants within each group showed similar averages for their respective categories. For males, the average values of height, weight, WC, BMI, and CRF did not differ significantly between the groups. The same pattern was held for female participants’ height. However, the CSR categories observed significant differences in female participants’ weight, WC, BMI, and CRF. Specifically, for females in the CSR < 1 group, the average weight was 60.5 ± 11.2 kg, significantly higher than those in the 1 ≤ CSR < 1.5 group (57.6 ± 9.1 kg). The WC was also larger in the CSR < 1 group (78.9 ± 10.6 cm) compared to the 1 ≤ CSR < 1.5 group (76.1 ± 9.1 cm). Similarly, the BMI was 23.7 ± 4.1 kg/m2 in the CSR < 1 group, which was significantly higher than the 1 ≤ CSR < 1.5 group (22.5 ± 3.4 kg/m2). The fasting glucose levels in the CSR < 1 group (95.7 ± 15.3 mg/dL) were significantly lower than the 1.5 ≤ CSR group (96.6 ± 18.0 mg/dL). The HbA1c levels were marginally lower in the 1 ≤ CSR < 1.5 group (5.5 ± 0.6%) compared to the 1.5 ≤ CSR group (5.6 ± 0.6%). For the entire participant group (combined male and female), those in the CSR < 1 group had an average weight of 66.5 ± 13.4 kg, significantly higher than the 1 ≤ CSR < 1.5 group (64.8 ± 13.9 kg). The WC was also larger in the CSR < 1 group (82.6 ± 10.8 cm) compared to the 1 ≤ CSR < 1.5 group (80.9 ± 10.8 cm). The BMI was 24.2 ± 3.8 kg/m2 in the CSR < 1 group, which was significantly higher than the 1 ≤ CSR < 1.5 group (23.5 ± 3.7 kg/m2). The HbA1c levels were marginally lower in the 1 ≤ CSR < 1.5 group (5.6 ± 0.7%) compared to the 1.5 ≤ CSR group (5.7 ± 0.8%).

**Table 1.**
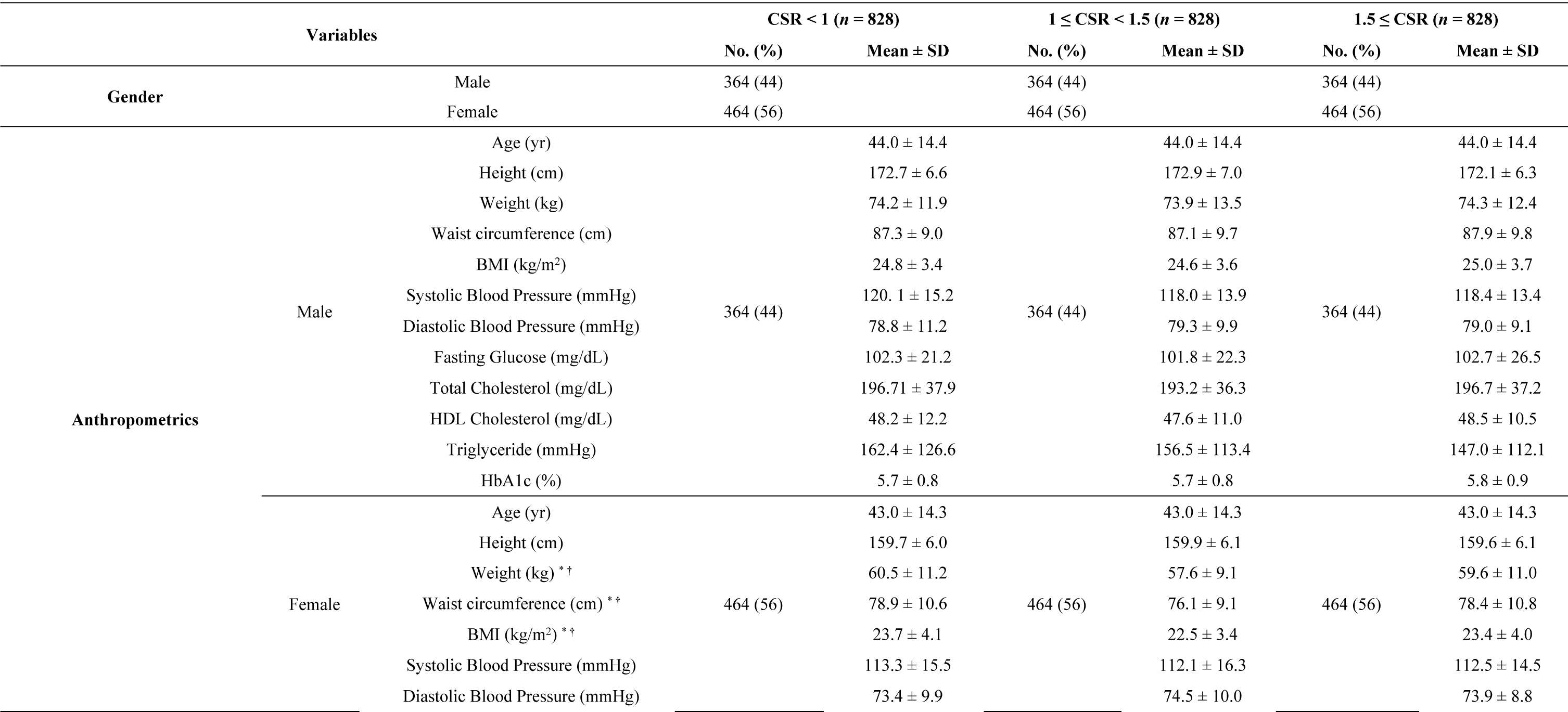
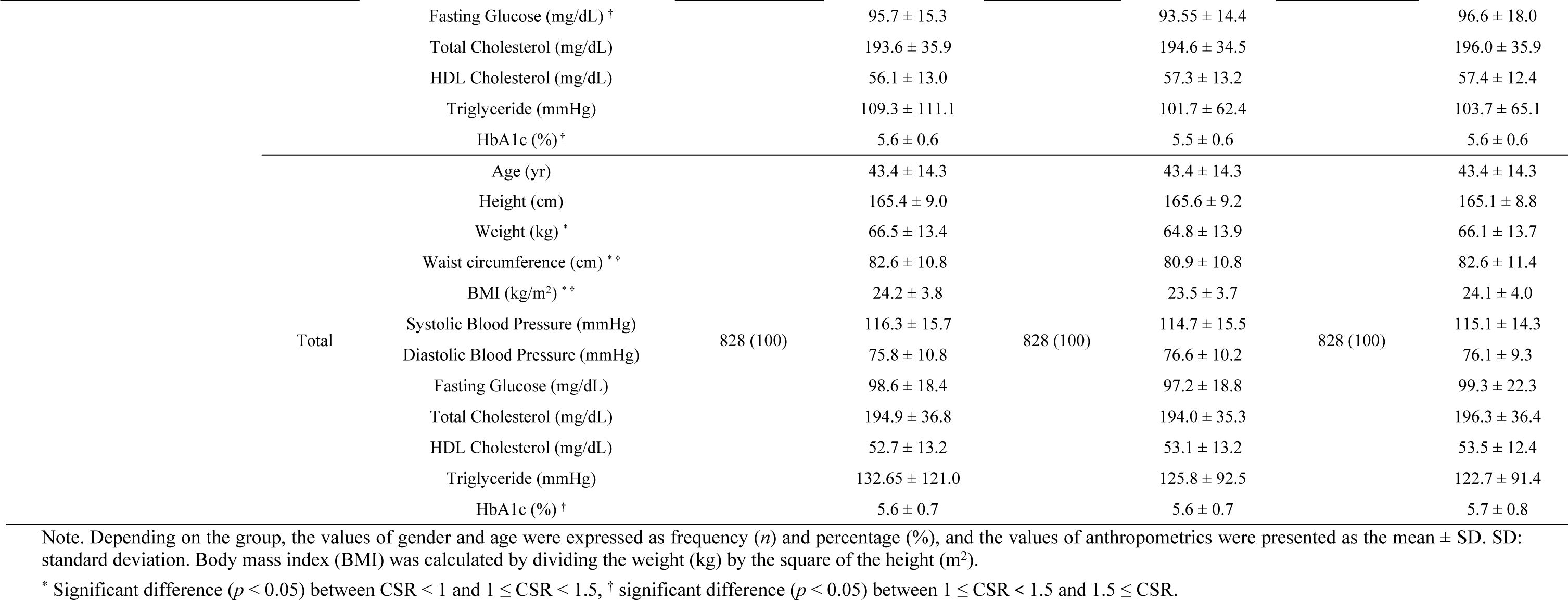
Characteristics and anthropometrics for participants in three groups (n = 2,484)

Table 2 shows the distribution of various demographic and health characteristics across three groups. There was a significant difference between the three CSR groups regarding their residence type (X2 = 32.70, *p* < 0.001). The majority of each group lived in apartments, but there was a slightly higher proportion of individuals living in single-family houses in the CSR < 1 group and a larger proportion living in “other” types of residence in the 1 ≤ CSR < 1.5 group. For the occupational status, the CSR groups significantly differed in their occupational status (X2 = 83.74, *p* < 0.001). The CSR < 1 group had a higher proportion of white-collar workers, while the 1 ≤ CSR < 1.5 and 1.5 ≤ CSR groups had a higher proportion of pink and blue-collar workers. The work schedule showed a significant difference (X2 = 9.87, *p* < 0.05), with day work being the most common in all groups. Income quintiles also differed significantly (X2 = 31.26, *p* < 0.001), with lower income quintiles more common in the CSR < 1 and 1.5 ≤ CSR groups, while the 1 ≤ CSR < 1.5 group had more high-income individuals. However, drinking and smoking statuses showed no significant differences between groups. Meeting guidelines for moderate PA showed a significant difference (X2 = 6.35, *p* < 0.05) but not for vigorous PA. The number of walking days per week (X2 = 35.87, *p* < 0.001) and SB time (X2 = 47.54, *p* < 0.001) significantly varied across the groups. Self-perceived body shape, health status, and stress levels categories also significantly differed between CSR groups. Among CRF, only fasting glucose and HDL cholesterol levels showed significant differences between the groups. In contrast, others, such as BMI, hypertension, total cholesterol, triglycerides, and HbA1c levels, showed no significant differences.

**Table 2.** Result of chi-square analysis examining differences in general characteristics across three groups (*n* = 2,484)

| Variables |  | CSR < 1 (%) | 1 ≤ CSR < 1.5 (%) | 1.5 ≤ CSR (%) | X<sup>2</sup> |
| --- | --- | --- | --- | --- | --- |
| Type of Residence | Single-family house | 241 (29.1) | 168 (20.3) | 219 (26.4) | 32.70 *** |
|  | Apartment | 473 (57.1) | 471 (56.9) | 466 (56.3) |  |
|  | Others | 114 (13.8) | 189 (22.8) | 143 (17.3) |  |
| Occupational Status | White | 306 (37.0) | 391 (47.2) | 383 (46.3) | 83.74 *** |
|  | Pink | 85 (10.3) | 70 (8.5) | 69 (8.3) |  |
|  | Blue | 197 (23.8) | 113 (13.6) | 227 (27.4) |  |
|  | Not classified | 240 (29.0) | 254 (30.7) | 149 (18.0) |  |
| Work Schedule | Day | 542 (65.5) | 545 (65.8) | 574 (69.3) | 9.87 * |
|  | Night | 98 (11.8) | 79 (9.5) | 97 (11.7) |  |
|  | Shiftwork (or others) | 188 (22.7) | 204 (24.6) | 157 (19.0) |  |
| Income Quintile (Individual) | Low | 157 (19.0) | 157 (19.0) | 121 (14.6) | 31.26 *** |
|  | Lower middle | 176 (21.3) | 152 (18.4) | 161 (19.4) |  |
|  | Middle | 168 (20.3) | 139 (16.8) | 177 (21.4) |  |
|  | Upper middle | 176 (21.3) | 156 (18.8) | 192 (23.2) |  |
|  | High | 151 (18.8) | 224 (27.1) | 177 (21.4) |  |
| Drinking Status | Abstained in the last year | 174 (21.0) | 152 (18.4) | 170 (20.5) | 7.96 |
|  | Once a month | 200 (24.2) | 188 (22.7) | 169 (20.4) |  |
|  | 2 – 4 times a month | 206 (24.9) | 229 (27.7) | 208 (25.1) |  |
|  | At least twice a week | 248 (30.0) | 259 (31.3) | 281 (33.9) |  |
| **Smoking Status** | Current smoker | 163 (19.7) | 194 (23.4) | 186 (22.5) | 4.39 |
|  | Ex-smoker | 171 (20.7) | 172 (20.8) | 178 (21.5) |  |
|  | Never smoked | 494 (59.7) | 462 (56.0) | 464 (55.8) |  |
| **Moderate PA** | Met | 359 (43.4) | 408 (49.3) | 371 (44.8) | 6.35 * |
|  | Not met | 469 (56.6) | 420 (50.7) | 457 (55.2) |  |
| **Vigorous PA** | Met | 85 (10.3) | 68 (8.2) | 74 (8.9) | 2.16 |
|  | Not met | 743 (89.7) | 760 (91.8) | 754 (91.1) |  |
| **Walking Days (wk)** | Not participating | 132 (15.9) | 95 (11.5) | 145 (17.5) | 35.87 *** |
|  | 1 – 3 days | 255 (30.8) | 203 (24.5) | 230 (27.8) |  |
|  | 4 – 6 days | 230 (27.8) | 237 (28.6) | 242 (29.2) |  |
|  | Everyday | 211 (25.5) | 293 (35.4) | 211 (25.5) |  |
| **SB time (min)** | < 420 | 324 (39.1) | 239 (28.9) | 228 (27.5) | 47.54 *** |
|  | 420 ≤ Time < 600 | 200 (24.2) | 213 (25.7) | 170 (20.5) |  |
|  | 600 ≤ | 304 (36.7) | 376 (45.4) | 430 (51.9) |  |
| **Self-perceived Body Shape** | Thin | 107 (12.9) | 116 (14.0) | 102 (12.3) | 12.86 * |
|  | Normal | 295 (35.6) | 352 (42.5) | 309 (37.3) |  |
|  | Obese | 426 (51.4) | 360 (43.5) | 417 (50.4) |  |
| **Self-perceived Health Status** | Good | 256 (30.9) | 297 (35.9) | 253 (30.6) | 10.61 * |
|  | Average | 427 (51.6) | 423 (51.1) | 438 (52.9) |  |
|  | Bad | 145 (17.5) | 108 (13.0) | 137 (16.5) |  |
| **Self-perceived Stress Level** | Not at all | 115 (13.9) | 107 (12.9) | 83 (10.0) | 24.74 *** |
|  | A little | 451 (54.5) | 490 (59.2) | 426 (51.4) |  |
|  | Quite | 262 (31.6) | 231 (27.9) | 319 (38.5) |  |
| **BMI (kg/m<sup>2</sup>)** | Underweight | 30 (3.6) | 48 (5.8) | 39 (4.7) | 15.22 * |
|  | Normal | 306 (37.0) | 350 (42.3) | 301 (36.4) |  |
|  | Overweight | 185 (22.3) | 178 (21.5) | 194 (23.4) |  |
|  | Obese | 307 (37.1) | 252 (30.4) | 294 (35.5) |  |
| **Hypertension (mmHg)** | Normal | 525 (63.4) | 556 (67.1) | 545 (65.8) | 5.77 |
|  | Prehypertension | 229 (27.7) | 208 (25.1) | 230 (27.8) |  |
|  | Hypertension | 74 (8.9) | 64 (7.7) | 53 (6.4) |  |
| **Fasting Glucose (mg/dL)** | Normal | 558 (67.4) | 611 (73.8) | 559 (67.5) | 11.73 * |
|  | Impaired glucose | 221 (26.7) | 170 (20.5) | 221 (26.7) |  |
|  | Diabetes | 49 (5.9) | 47 (5.7) | 48 (5.8) |  |
| **Total Cholesterol (mg/dL)** | Normal | 486 (58.7) | 490 (59.2) | 461 (55.7) | 3.81 |
|  | Borderline | 248 (30.0) | 258 (31.2) | 276 (33.3) |  |
|  | Hyperlipidemia | 94 (11.4) | 80 (9.7) | 91 (11.0) |  |
| **HDL Cholesterol (mg/dL)** | Low | 126 (15.2) | 121 (14.6) | 87 (10.5) | 10.75 * |
|  | Normal | 478 (57.7) | 470 (56.8) | 514 (62.1) |  |
|  | High | 224 (27.1) | 237 (28.6) | 227 (27.4) |  |
| **Triglyceride (mmHg)** | Normal | 593 (71.6) | 612 (73.9) | 632 (76.3) | 4.84 |
|  | Borderline | 115 (13.9) | 104 (12.6) | 97 (11.7) |  |
|  | Danger | 120 (14.5) | 112 (13.5) | 99 (12.0) |  |
| **HbA1c (%)** | Normal | 609 (73.6) | 643 (77.7) | 601 (72.6) | 6.63 |
|  | Borderline | 154 (18.6) | 130 (15.7) | 155 (18.7) |  |
|  | Diabetes | 65 (7.9) | 55 (6.6) | 72 (8.7) |  |

Note. The values are presented as n (%). \*\*\* $p < 0.001$ , \*\* $p < 0.01$ , \* $p < 0.05$ , PA: Physical Activity, SB: Sedentary Behavior

Table 3 presents the results of an ANOVA analysis for different variables across three groups. Firstly, for moderate PA, no significant difference was observed (p = 0.91), with mean values of 191.3 ± 293.2, 187.6 ± 238.0, and 193.2 ± 288.7 for the CSR < 1.0, 1.0 ≤ CSR < 1.5, and 1.5 ≤ CSR groups respectively. However, vigorous PA displayed a significant mean difference between CSR < 1.0 (30.6 ± 133.6) and the 1.0 ≤ CSR < 1.5 (17.4 ± 63.8) groups (p = 0.04). SB time revealed significant mean differences between all groups (p < 0.001): CSR < 1.0 (482.1 ± 209.9), 1.0 ≤ CSR < 1.5 (520.6 ± 202.9), and 1.5 ≤ CSR (555.8 ± 227.7). Both sleep duration on weekdays and weekends demonstrated significant mean differences among all groups (p < 0.001), with respective values for CSR < 1.0 of 458.9 ± 75.8 (weekday) and 388.5 ± 81.0 (weekend), for 1.0 ≤ CSR < 1.5 of 422.0 ± 70.4 (weekday) and 466.5 ± 82.5 (weekend), and for 1.5 ≤ CSR of 327.3 ± 68.1 (weekday) and 562.2 ± 107.3 (weekend). Weekly Sleep Duration also exhibited significant mean differences (p < 0.001) between CSR < 1.0 (423.7 ± 75.1) and both 1.0 ≤ CSR < 1.5 (444.2 ± 72.3) and 1.5≤CSR (444.7 ± 81.2), but not between 1.0 ≤ CSR < 1.5 and 1.5≤CSR. Lastly, worktime showed significant mean differences among all groups (p < 0.001): CSR < 1.0 (1833.1 ± 1204.2), 1.0 ≤ CSR < 1.5 (1858.7 ± 1260.7), and 1.5 ≤ CSR (2270.9 ± 1157.7).

**Table 3.**
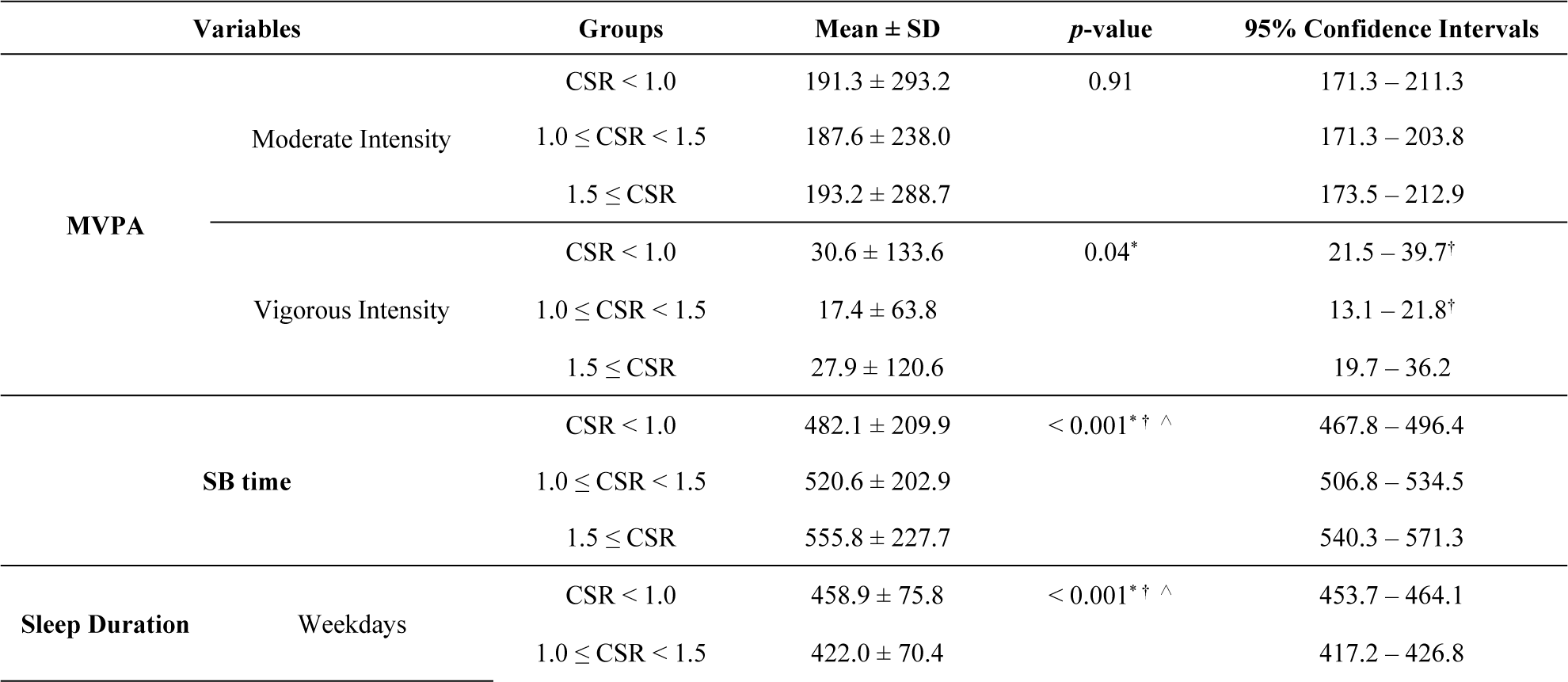
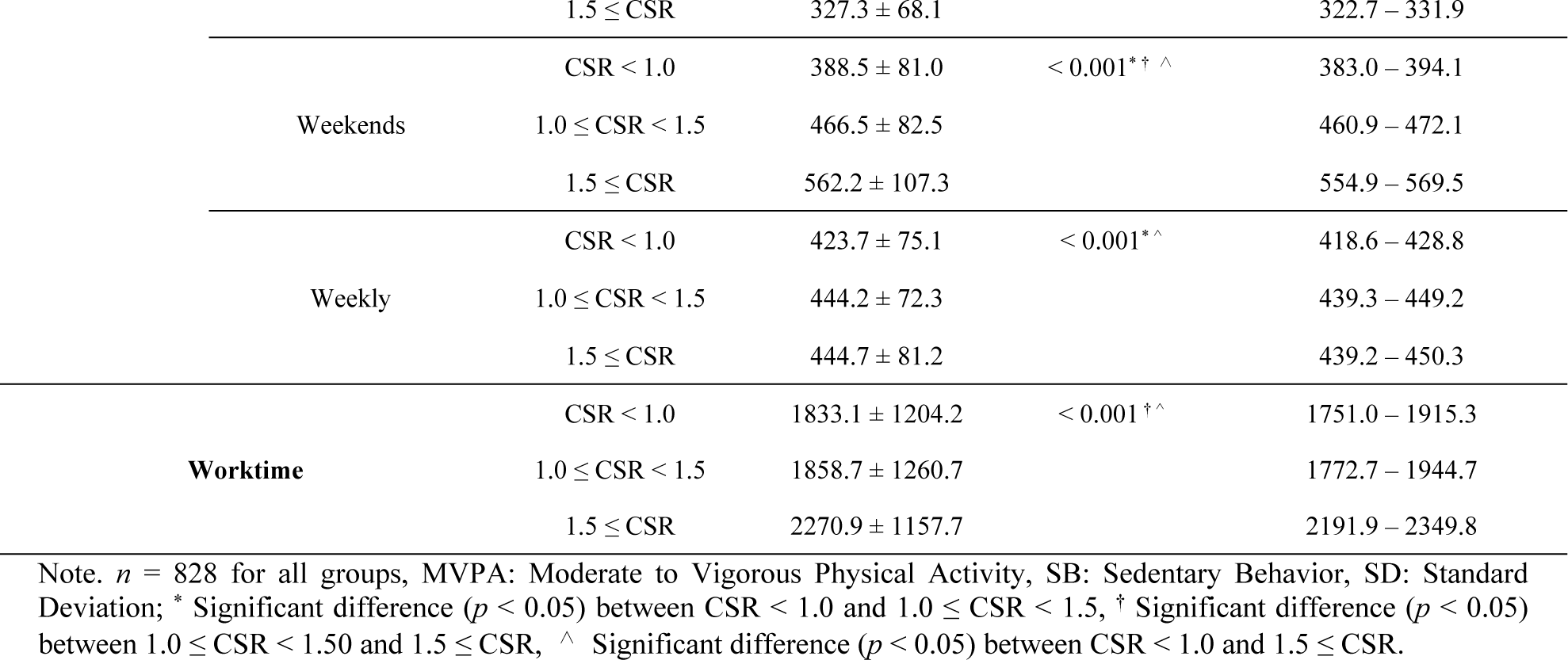
Comparative ANOVA analysis of health-related factors across CSR groups (n = 2,484)

Table 4 shows the multinomial logistic regression results with 1.0 ≤ CSR < 1.5 as the reference group. For the ‘Type of Residence’, those residing in a ‘Single-family house’ were 2.42 times more likely to be in the CSR < 1 group (OR = 2.42, 95% CI = 1.77 – 3.32, *p*-value < 0.001) and 1.78 times more likely to be in the CSR ≥ 1.5 group (OR = 1.78, 95% CI = 1.3 – 2.43, *p*-value < 0.001). Similarly, those living in an “Apartment” were 2.01 times more likely to be in the CSR < 1 group (OR = 2.01, 95% CI = 1.52 – 2.67, *p*-value < 0.001) and 1.48 times more likely to be in the CSR ≥ 1.5 group (OR = 1.48, 95% CI = 1.13 – 1.95, *p*-value = 0.004). In the ‘Occupational Status’, compared to the ‘Not classified’ reference group, the “White” workers were 0.71 times less likely to be in the CSR < 1 group (OR = 0.71, 95% CI = 0.52 – 0.97, *p*-value = 0.03) but 1.71 more likely to be in the CSR ≥ 1.5 group (OR = 1.71, 95% CI = 1.22 – 2.39, *p*-value = 0.002). The “Pink” workers were significantly more likely to be in the CSR ≥ 1.5 group (OR = 3.93, 95% CI = 2.7 – 5.71, *p*-value < 0.001). The “Blue” workers showed 1.76 times more likely to be in the CSR ≥ 1.5 group (OR = 1.76, 95% CI = 1.12 – 2.77, *p*-value = 0.01). In the ‘Work Schedule’, people working in shiftwork or other work schedules were 0.8 times as likely to be in the CSR < 1 group (OR = 0.8, 95% CI = 0.58 – 1.09, *p*-value = 0.15). However, they were 1.13 times more likely to be in the CSR ≥ 1.5 group (OR = 1.13, 95% CI = 0.82 – 1.57, *p*-value = 0.46). Night shift workers were 1.27 times as likely to be in the CSR < 1 group (OR = 1.27, 95% CI = 0.91 – 1.79, *p*-value = 0.16) and 1.29 times as likely to be in the CSR ≥ 1.5 group (OR = 1.29, 95% CI = 0.91 – 1.81, *p*-value = 0.15). For the ‘Income Quintile (individual)’, those in the “low” category were 0.86 times as likely to be in the CSR < 1 group (OR = 0.86, 95% CI = 0.62 – 1.2, *p*-value = 0.37) and 0.63 times as likely to be in the CSR ≥ 1.5 group (OR = 0.63, 95% CI = 0.45 – 0.89, *p*-value = 0.008). The ‘lower middle’, ‘upper middle’, and ‘high’ categories showed no statistically significant differences in the CSR < 1 group. However, those in the “high” category were 0.71 times as likely to be in the CSR ≥ 1.5 group (OR = 0.71, 95% CI = 0.52 – 0.98, *p*-value = 0.04). Regarding the ‘Drinking Status’, people who drank once a month were 0.63 times as likely to be in the CSR ≥ 1.5 group (OR = 0.63, 95% CI = 0.46 – 0.89, *p*-value = 0.007). Those who drank 2 – 4 times a month were 0.75 times as likely to be in the CSR ≥ 1.5 group (OR = 0.75, 95% CI = 0.55 – 1.02, *p*-value = 0.07). In terms of ‘Smoking Status’, current smokers were 0.79 times as likely to be in the CSR < 1 group (OR = 0.79, 95% CI = 0.61 – 1.01, *p*-value = 0.06).

**Table 4.** Multinomial logistic regression results (*n* = 2,484)

| Variables |  | CSR < 1 |  |  |  | 1.5 ≤ CSR |  |  |  |
| --- | --- | --- | --- | --- | --- | --- | --- | --- | --- |
|  |  | 95% CI |  |  |  | 95% CI |  |  |  |
|  |  | *p*-value | OR | Lower | Upper | *p*-value | OR | Lower | Upper |
| Type of Residence | Others |  | Ref. |  |  |  | Ref. |  |  |
|  | Single-family house | < 0.001 | 2.42*** | 1.77 | 3.32 | < 0.001 | 1.78*** | 1.30 | 2.43 |
|  | Apartment | < 0.001 | 2.01*** | 1.52 | 2.67 | 0.004 | 1.48** | 1.13 | 1.95 |
| Occupational Status | Not classified |  | Ref. |  |  |  | Ref. |  |  |
|  | White | 0.03 | 0.71* | 0.52 | 0.97 | 0.002 | 1.71** | 1.22 | 2.39 |
|  | Pink | 0.06 | 1.42 | 0.99 | 2.02 | < 0.001 | 3.93*** | 2.70 | 5.71 |
|  | Blue | 0.89 | 0.97 | 0.63 | 1.48 | 0.01 | 1.76* | 1.12 | 2.77 |
| Work Schedule | Day |  | Ref. |  |  |  | Ref. |  |  |
|  | Shiftwork (or others) | 0.15 | 0.80 | 0.58 | 1.09 | 0.46 | 1.13 | 0.82 | 1.57 |
|  | Night | 0.16 | 1.27 | 0.91 | 1.79 | 0.15 | 1.29 | 0.91 | 1.81 |
| Income Quintile (Individual) | Middle |  | Ref. |  |  |  | Ref. |  |  |
|  | Low | 0.37 | 0.86 | 0.62 | 1.20 | 0.008 | 0.63** | 0.45 | 0.89 |
|  | Lower middle | 0.80 | 1.04 | 0.75 | 1.44 | 0.45 | 0.88 | 0.63 | 1.22 |
|  | Upper middle | 0.85 | 1.03 | 0.75 | 1.43 | 0.93 | 1.02 | 0.74 | 1.40 |
|  | High | 0.02 | 0.68* | 0.49 | 0.94 | 0.04 | 0.71* | 0.52 | 0.98 |
| Drinking Status | Abstained in the last year |  | Ref. |  |  |  | Ref. |  |  |
|  | Once a month | 0.30 | 0.84 | 0.61 | 1.17 | 0.007 | 0.63** | 0.46 | 0.89 |
|  | 2 – 4 times a month | 0.26 | 0.84 | 0.62 | 1.14 | 0.07 | 0.75 | 0.55 | 1.02 |
|  | At least twice a week | 0.4 | 0.88 | 0.66 | 1.18 | 0.51 | 0.91 | 0.67 | 1.21 |
| Smoking Status | Never smoked |  | Ref. |  |  |  | Ref. |  |  |
|  | Current smoker | 0.06 | 0.79 | 0.61 | 1.01 | 0.88 | 0.98 | 0.76 | 1.26 |
|  | Ex-smoker | 0.69 | 0.95 | 0.73 | 1.23 | 0.78 | 1.04 | 0.80 | 1.34 |
| Moderate PA | Not met |  | Ref. |  |  |  | Ref. |  |  |
|  | Met | 0.50 | 0.92 | 0.73 | 1.16 | 0.52 | 1.08 | 0.86 | 1.36 |
| Vigorous PA | Not met |  | Ref. |  |  |  | Ref. |  |  |
|  | Met | 0.003 | 1.71** | 1.20 | 2.46 | 0.12 | 1.34 | 0.92 | 1.94 |
| Walking Days (wk) | Not participating |  | Ref. |  |  |  | Ref. |  |  |
|  | 1~3 days | 0.92 | 1.02 | 0.73 | 1.42 | 0.26 | 0.82 | 0.59 | 1.15 |
|  | 4~6 days | 0.13 | 0.76 | 0.53 | 1.08 | 0.04 | 0.69* | 0.49 | 0.98 |
|  | Everyday | 0.003 | 0.58** | 0.41 | 0.83 | < 0.001 | 0.53*** | 0.37 | 0.75 |
| SB Time (min) | < 420 |  | Ref. |  |  |  | Ref. |  |  |
|  | 420 ≤ Time < 600 | 0.02 | 0.72* | 0.55 | 0.95 | 0.62 | 0.93 | 0.70 | 1.24 |
|  | 600 ≤ | 0.001 | 0.66** | 0.51 | 0.84 | 0.006 | 1.42** | 1.11 | 1.82 |
| Self-perceived Body Shape | Normal |  | Ref. |  |  |  | Ref. |  |  |
|  | Thin | 0.14 | 1.30 | 0.92 | 1.85 | 0.99 | 1.00 | 0.70 | 1.42 |
|  | Obese | 0.048 | 1.33* | 1.00 | 1.76 | 0.14 | 1.24 | 0.93 | 1.64 |
| Self-perceived Health Status | Average |  | Ref. |  |  |  | Ref. |  |  |
|  | Good | 0.53 | 0.93 | 0.74 | 1.17 | 0.42 | 0.91 | 0.72 | 1.15 |
|  | Bad | 0.22 | 1.21 | 0.89 | 1.63 | 0.65 | 1.07 | 0.79 | 1.46 |
| Self-perceived Stress Level | Not at all |  | Ref. |  |  |  | Ref. |  |  |
|  | A little | 0.31 | 0.85 | 0.62 | 1.16 | 0.73 | 1.06 | 0.76 | 1.48 |
|  | Quite | 0.78 | 1.05 | 0.74 | 1.48 | 0.003 | 1.72** | 1.20 | 2.47 |
| BMI (kg/m2) | Normal |  | Ref. |  |  |  | Ref. |  |  |
|  | Underweight | 0.11 | 0.65 | 0.38 | 1.11 | 0.89 | 1.04 | 0.62 | 1.73 |
|  | Overweight | 0.47 | 1.12 | 0.83 | 1.52 | 0.41 | 1.14 | 0.84 | 1.54 |
|  | Obese | 0.34 | 1.18 | 0.84 | 1.67 | 0.56 | 1.11 | 0.78 | 1.57 |
| Hypertension (mmHg) | Normal |  | Ref. |  |  |  | Ref. |  |  |
|  | Prehypertension | 0.57 | 0.93 | 0.73 | 1.19 | 0.81 | 0.97 | 0.76 | 1.24 |
|  | Hypertension | 0.65 | 0.91 | 0.62 | 1.35 | 0.08 | 0.69 | 0.45 | 1.04 |
| Fasting Glucose (mg/dL) | Normal |  | Ref. |  |  |  | Ref. |  |  |
|  | Impaired glucose | 0.09 | 1.26 | 0.96 | 1.66 | 0.06 | 1.31 | 0.99 | 1.72 |
|  | Diabetes | 0.60 | 0.82 | 0.40 | 1.71 | 0.34 | 0.70 | 0.34 | 1.46 |
| Total Cholesterol (mg/dL) | Normal |  | Ref. |  |  |  | Ref. |  |  |
|  | Borderline | 0.17 | 0.85 | 0.68 | 1.07 | 0.69 | 1.05 | 0.83 | 1.32 |
|  | Hyperlipidemia | 0.70 | 1.07 | 0.75 | 1.53 | 0.48 | 1.14 | 0.80 | 1.63 |
| HDL Cholesterol (mg/dL) | Desirable |  | Ref. |  |  |  | Ref. |  |  |
|  | Low | 0.53 | 0.90 | 0.66 | 1.24 | 0.007 | 0.62** | 0.44 | 0.88 |
|  | High | 0.43 | 1.1 | 0.86 | 1.41 | 0.84 | 1.03 | 0.80 | 1.31 |
| Triglyceride (mmHg) | Normal |  | Ref. |  |  |  | Ref. |  |  |
|  | Borderline | 0.95 | 0.99 | 0.72 | 1.36 | 0.13 | 0.77 | 0.55 | 1.08 |
|  | Danger | 0.80 | 1.05 | 0.74 | 1.48 | 0.37 | 0.85 | 0.59 | 1.21 |
| HbA1c (%) | Normal |  | Ref. |  |  |  | Ref. |  |  |
|  | Borderline | 0.86 | 0.97 | 0.72 | 1.31 | 0.47 | 1.12 | 0.83 | 1.51 |
|  | Diabetes | 0.56 | 1.22 | 0.63 | 2.36 | 0.06 | 1.87 | 0.97 | 3.60 |

Note. \*\*\* $p < 0.001$ , \*\* $p < 0.01$ , \* $p < 0.05$ , S.E: Standard Error, OR: Odd Ratio, CI: Confidence Interval, PA: Physical Activity, SB: Sedentary Behavior, the reference group is the $1.0 \leq \text{CSR} < 1.5$ group.

In terms of ‘Moderate PA’, individuals who met the WHO’s guidelines were 0.92 times as likely to be in the CSR < 1 group (OR = 0.92, 95% CI = 0.73 – 1.16, *p*-value = 0.5), and were 1.08 times as likely to be in the CSR ≥ 1.5 group (OR = 1.08, 95% CI = 0.86 – 1.36, *p*-value = 0.52). For ‘Vigorous PA’, those who met the criteria were 1.71 times more likely to be in the CSR < 1 group (OR = 1.71, 95% CI = 1.2 – 2.46, *p*-value = 0.003). However, they were 1.34 times more likely to be in the CSR ≥ 1.5 group (OR = 1.34, 95% CI = 0.92 – 1.94, *p*-value = 0.12). For ‘Walking Days per week’, those who walked every day were less likely to either CSR group compared to those who didn’t participate at all, being 0.58 times in the CSR < 1 group (OR = 0.58, 95% CI = 0.4 – 0.83, *p*-value = 0.003) and 0.53 times in the CSR ≥ 1.5 group (OR = 0.53, 95% CI = 0.37 – 0.75, *p*-value < 0.001). Those who walked 4 – 6 days per week were 0.76 times as likely to be in the CSR < 1 group (OR = 0.76, 95% CI = 0.53 – 1.08, *p*-value = 0.13) and 0.69 times as likely to be in the CSR ≥ 1.5 group (OR = 0.69, 95% CI = 0.49 – 0.98, *p*-value = 0.04). Those who walked 1 – 3 days per week were just as likely to be in either CSR group as non-participants. In terms of ‘SB Time’, those with a time between 420 – 600 minutes were 0.72 times as likely to be in the CSR < 1 group (OR = 0.72, 95% CI = 0.55 – 0.95, *p*-value = 0.02) and were just as likely to be in the CSR ≥ 1.5 group (OR = 0.93, 95% CI = 0.7 – 1.24, *p*-value = 0.62). Those with a time of 600 minutes or more were 0.66 times as likely to be in the CSR < 1 group (OR = 0.66, 95% CI = 0.51 – 0.84, *p*-value = 0.001), and were 1.42 times as likely to be in the CSR ≥ 1.5 group (OR = 1.42, 95% CI = 1.11 – 1.82, *p*-value = 0.006).

For ‘Self-perceived Body Shape’, those who perceived themselves as thin were 1.3 times as likely to be in the CSR < 1 group (OR = 1.3, 95% CI = 0.92 – 1.85, *p*-value = 0.14) compared to those who perceived themselves as normal. Individuals who perceived themselves as obese were 1.33 times as likely to be in the CSR < 1 group (OR = 1.33, 95% CI = 1 – 1.76, *p*-value = 0.048), and were 1.24 times as likely to be in the CSR ≥ 1.5 group (OR = 1.24, 95% CI = 0.93 – 1.64, *p*-value = 0.14). Regarding ‘Self-perceived Health Status’, those who perceived their health as good were 0.93 times as likely to be in the CSR < 1 group (OR = 0.93, 95% CI = 0.74 – 1.17, *p*-value = 0.53), and were 0.91 times as likely to be in the CSR ≥ 1.5 group (OR = 0.91, 95% CI = 0.72 – 1.15, *p*-value = 0.42). Those who perceived their health as bad were 1.2 times as likely to be in the CSR < 1 group (OR = 1.2, 95% CI = 0.89 – 1.63, *p*-value = 0.22). In terms of ‘Self-perceived Stress Level’, those who felt a little stressed were 0.85 times as likely to be in the CSR < 1 group (OR = 0.85, 95% CI = 0.62 – 1.16, *p*-value = 0.31). Those who felt quite stressed were 1.72 times as likely to be in the CSR ≥ 1.5 group (OR = 1.72, 95% CI = 1.2 – 2.47, *p*-value = 0.003).

Regarding ‘BMI’, individuals classified as underweight were 0.65 times as likely to be in the CSR < 1 group compared to normal-weight individuals (OR = 0.65, 95% CI = 0.38 – 1.11, *p*-value = 0.11). Obese individuals were 1.18 times as likely to be in the CSR < 1 group (OR = 1.18, 95% CI = 0.84 – 1.67, *p*-value = 0.34) and 1.11 times as likely to be in the CSR ≥ 1.5 group (OR = 1.11, 95% CI = 0.78 – 1.57, *p*-value = 0.56). In terms of ‘Hypertension’, those with hypertension were similarly likely to be in the CSR < 1 (OR = 0.91, 95% CI = 0.62 – 1.35, *p*-value = 0.65) and CSR ≥ 1.5 groups (OR = 0.69, 95% CI = 0.45 – 1.04, *p*-value = 0.08). For ‘Fasting Glucose’, individuals with impaired glucose were 1.26 times as likely to be in the CSR < 1 group (OR = 1.26, 95% CI = 0.96 – 1.66, *p*-value = 0.09) and 1.31 times as likely to be in the CSR ≥ 1.5 group (OR = 1.31, 95% CI = 0.99 – 1.72, *p*-value = 0.06). Regarding ‘Total Cholesterol’, individuals with borderline cholesterol were 0.85 times as likely to be in the CSR < 1 group (OR = 0.85, 95% CI = 0.68 – 1.07, *p*-value = 0.17) and were similarly likely to be in the CSR ≥ 1.5 group (OR = 1.05, 95% CI = 0.83 – 1.32, *p*-value = 0.69). Individuals with hyperlipidemia were 1.14 times as likely to be in the CSR ≥ 1.5 group (OR = 1.14, 95% CI = 0.80 – 1.63, *p*-value = 0.48). For ‘HDL Cholesterol’, individuals with low HDL were 0.62 times as likely to be in the CSR ≥ 1.5 group (OR = 0.62, 95% CI = 0.44 – 0.88, *p*-value < 0.01). In terms of ‘Triglyceride levels’, individuals with borderline levels were 0.77 times as likely to be in the CSR ≥ 1.5 groups (OR = 0.77, 95% CI = 0.55 – 1.08, *p*-value = 0.13) compared to those with normal levels. Lastly, for ‘HbA1c levels’’, individuals with borderline levels were 1.12 times as likely to be in the CSR ≥ 1.5 group (OR = 1.12, 95% CI = 0.83 – 1.51, *p*-value = 0.47). Those with diabetes were 1.22 times as likely to be in the CSR < 1 group (OR = 1.22, 95% CI = 0.63 – 2.36, *p*-value = 0.56) and 1.87 times as likely to be in the CSR ≥ 1.5 group (OR = 1.87, 95% CI = 0.97 – 3.60, *p*-value = 0.06).

## Discussion

Sleep disorders and sleep deprivation are becoming increasingly prevalent across the globe due to contemporary lifestyles. A common practice among Koreans is to catch-up sleep during weekends or days off to compensate for the sleep deficit accumulated over the work week. Such inconsistent sleep schedules and interruptions often lead to various health problems, ranging from decreased PA to increased mental health risks. These issues diminish an individual’s quality of life and contribute to broader clinical and public health challenges. Therefore, we examined the association between CSR of adults and health risk factors. In the present study, we demonstrated that the gap in CSR had statistically significant associations with PA levels, SB time, mental health, and CRF.

Our study reveals distinct differences in the anthropometric measures of participants based on the CSR groups, emphasizing a gender-specific impact. Although male participants showed no significant difference across CSR groups, we observed a downward trend in weight, WC, and BMI as CSR increased in female participants. Despite having higher weight-related measures, females in the CSR < 1 group exhibited significantly lower fasting glucose levels, indicating potential complex metabolic interplays. A marginal difference in HbA1c levels among the entire participant group (combined male and female) suggests a possible relation between better glycemic control and lower CSR. These findings suggest that CSR patterns might affect anthropometric characteristics differently based on gender, indicating a need for further research to elucidate the underlying mechanisms.

The type of residence significantly influenced CSR status. People who tended to get more sleep during weekdays and those who slept heavily on weekends were more likely to live in single-family houses or apartments. This implies sleep patterns and behaviors may impact the type of residence. Regarding occupational status, those who slept more on weekdays than weekends were less likely to be ‘White’ workers, but those who slept more on weekends than weekdays were more likely to belong to this job category. In contrast, people with high CSR were significantly more likely to be ‘Pink’ and ‘Blue’ workers. While the relation between low CSR group and work schedules was not clearly defined, those with high CSR group showed a slight tendency to belong to non-traditional schedules, such as shift work or night work. These results indicate that various occupational characteristics and work schedules can disrupt standard sleep patterns, suggesting that different work conditions could impact one’s sleep. The relationship between participants’ work time and CSR demonstrated significant variations across all groups, suggesting a correlation between longer work hours and increased CSR. This was especially evident in the group that sleeps more on weekends than on weekdays, where the correlation appeared more clearly. This aligns with Dahlgren and his colleagues (Dahlgren et al., 2006), which showed that individuals working long hours tend to sleep more on non-working days, likely to recover from sleep deprivation experienced during working days. CSR status also significantly influenced individuals’ income, with the high CSR group less likely to belong to the lower- and higher-income individuals. This means that people with high CSR are more likely to have very little or very high income. Regarding lifestyle habits, the high CSR group was less likely to belong to less frequent drinkers. People with high CSR had lower rates of drinking. This suggests that weekday and weekend sleep patterns can affect drinking rates and that drinking habits effectively prevent excessive CSR. Meanwhile, participants who slept more during the weekdays than on weekends were less likely to belong to current smokers than those with 1.0 ≤ CSR < 1.5. This might indicate that less sleep time during the weekdays than weekends is associated with a healthier lifestyle, such as not smoking. But additional research would be required to investigate the potential causal relationships between sleep patterns and smoking habits.

The relationship between CSR and adherence to WHO’s PA guidelines appears to differ based on the intensity of the activity. Individuals with a low CSR were less likely to meet the WHO’s recommendations for moderate-intensity PA, while those with a high CSR were more likely to do so. This suggests sleep behavior changes between weekdays and weekends may not substantially impact moderate PA. However, the low and high CSR groups were more likely to meet the WHO’s recommendations regarding vigorous-intensity PA than those with a typical sleeping pattern. These findings suggest a complex interplay between sleep behaviors and PA levels. The observed discrepancies in adherence to WHO’s PA guidelines among those with different CSR levels imply that sleep patterns may influence how individuals adjust their PA intensity. Interestingly, for ‘Walking Days per week’, our findings suggest an association between regular walking and a more consistent sleep pattern throughout the week. This could indicate that irregular sleep patterns, either sleeping more on weekdays or having large amounts of sleep on weekends (presumably to catch up on sleep debt accrued during the week), might be associated with lower levels of regular PA, such as walking. Furthermore, those who sleep more on weekdays might miss out on morning or daytime opportunities for walking due to their extended sleep schedule. Conversely, individuals sleeping heavily on weekends might use this time to recover from sleep debt accumulated over the week, limiting their time or energy for PA. In terms of ‘SB Time’, A lower CSR, typically observed in participants who slept more during the weekdays, was associated with reduced SB time, specifically in those spending between 420 – 600 minutes or more than 600 minutes in SB. This suggests less sleep debt during weekdays may encourage a more active lifestyle with lower SB time. On the other hand, a higher CSR was more prevalent among participants with a daily SB time of 600 minutes or more. This suggests a lifestyle with more SB time might lead to greater sleep debt during the working week, necessitating more catch-up sleep and a higher CSR. These findings highlight the complex relationship between catch-up sleep needs and a sedentary lifestyle. We suggest mitigating sleep debt could be critical in reducing SB time.

In examining ‘Self-perceived Body Shape’, individuals who slept more during weekdays tend to identify as thin or obese. However, those who had more sleep time during weekends showed a specific tendency to perceive themselves as obese. The study indicates an association between sleeping more during weekdays, catching up on sleep excessively during weekends, and the self-perception of being obese. Considering the relationship between self-perception of body shapes and mental health conditions, such as depression and anxiety, we suggest that sleep hygiene might directly connect to the mental health (Darimont et al., 2020). Participants tended to sleep more on weekdays, and those who tended to compensate by sleeping excessively on the weekend showed a slightly decreased possibility of perceiving their health as ‘good’. These findings suggest a potential relationship between sleep patterns and self-perception of health. Specifically, deviations from an average sleep pattern appear to be associated with a less positive perception of one’s health. These results emphasize the importance of maintaining a balanced and consistent sleep routine for overall health and well-being. Regarding stress levels, it was observed that participants who slept excessively on weekends compared to weekdays were slightly more likely to consider themselves a little stressed. However, there was a significant difference between the groups in answers to the questionnaire that felt that the stress level was ‘quite’. This suggests a direct relationship between longer sleep duration on weekends and stress relief. However, the increase in feeling ‘very’ stressed out among these participants means there may be a tipping point where excessive sleep can lead to increased stress. Meanwhile, the pattern observed among people who sleep a lot on weekends may be due to a phenomenon known as ‘social jet lag’. It refers to a discrepancy between an individual’s biological clock and social schedule, often caused by sleeping late or staying up late on weekends. You may get more sleep, but it can disrupt your body’s circadian rhythm, heightening stress.

Among the CRF, BMI shows a higher tendency towards obesity in individuals who slept more on weekdays and those who slept excessively on weekends than on weekdays. Individuals who slept more on weekdays were less likely to be underweight. This pattern may emphasize the complex interplay between obesity, lifestyle behaviors, and sleep duration. Consistent sleep patterns also help maintain the balance of the hunger and satiety-regulating hormones leptin and ghrelin, which can contribute to a healthier BMI (Taheri et al., 2004). Leptin signals the brain to suppress appetite, and ghrelin, which stimulates appetite, can be negatively impacted by irregular sleep (Van Cauter et al., 2008). These findings underline the need to improve sleep quality and consistency. In addition, groups had similarly diverse differences in other variables related to CRF. First, in the case of low CSR compared to the group with regular sleep patterns, the possibility of belonging to prehypertension and hypertension was low, and the probability of belonging to impaired glucose with fasting glucose was high, but a lower propensity towards diabetes. Total cholesterol, triglyceride, and HbA1c were less likely to belong to the borderline, but conversely, hyperlipidemia, danger, and diabetes were high. Paradoxically, the likelihood of them having hyperlipidemia and diabetes was higher, and their overall health risk was also elevated. A noticeable difference appeared in the group with high CSR. While they showed a similar pattern as the low CSR group regarding hypertension, fasting glucose, and HDL cholesterol, the odds ratios were more pronounced. The highest odds ratio was found with HbA1c, indicating that those catch-up sleep excessively over weekends were about twice as likely to have diabetes as those with regular sleep patterns. Contrary to our expectations that irregular sleep patterns would result in negative CRF, our results indicated a decreased risk for cardiometabolic diseases relating to hypertension, fasting glucose, and triglycerides. This suggests that managing blood pressure and fasting blood sugar is multifactorial and not confined to sleep patterns. Hence, while sleep is a vital aspect of overall health, it forms only one facet of a comprehensive approach to maintaining a healthy body. This approach includes a balanced diet, regular physical activity, stress management, and avoiding harmful habits such as smoking and excessive alcohol consumption. Consequently, our study underscores the complexity of maintaining physical health and the significance of adopting a holistic perspective toward health and lifestyle.

This study revealed several positive strengths. First, the strength of this study is the matching of gender and age using a nationally representative data set. In addition, this is the first study to investigate the associations between CSR and various factors such as socioeconomic status, PA level, SB time, self-perception level, and cardiometabolic risk factors. As far as we know, no prior research has previously delved into such a diverse range of aspects concerning CSR. This positions our study at the forefront of this area of research, shedding new light on the complex interconnections between lifestyle, health, and sleep habits. However, this study has a few limitations. The cross-sectional design of our study limits our ability to conclude the causality or long-term dynamics of these relationships. Additionally, the potential for bias due to the reliance on participants’ self-reported data cannot be discounted. Therefore, we suggest that future studies should strive to expand upon these findings by delineating the causal links between CSR and the array of factors analyzed in this research. A transition to a longitudinal study design, alongside using objective sleep and physical activity measurements, could afford more profound and precise insights into these relationships. Importantly, subsequent investigations should strive to shed light on the specific mechanisms whereby these elements shape sleep behaviors and metabolic health. This could ultimately facilitate better health outcomes and improved quality of life for these individuals.

## Conclusion

In conclusion, our research indicates that weekend CSR can provide significant insights into various health behaviors, lifestyle habits, and CRF. The modern lifestyle, heavily influenced by widespread technology use, can drastically alter our sleep patterns, substantially impacting our physical and mental health. Shifts in CSR could signal changes in PA levels, SB duration, and other health indicators, ultimately shaping overall well-being. Therefore, a comprehensive health management approach incorporating CSR considerations becomes increasingly essential. Government agencies, healthcare providers, and individuals should acknowledge the significance of sleep patterns on health and incorporate these insights when devising health promotion strategies. The differences among the groups observed in our study highlight the necessity for more detailed investigations into sleep patterns, enhancing our understanding of their impact on adult health. This knowledge can aid in creating more targeted and effective health interventions for individuals across different strata of society.

## Data Availability

The data underlying the results presented in the study are available in the [Korea National Health and Nutrition Examination Survey] repository, [https://knhanes.kdca.go.kr/knhanes/sub03/sub03_02_05.do]

## Acknowledgments

We want to thank for participants who took part in our experiments.

